# Antimicrobial Resistance, Evidences on Irrational Anti-microbial Prescribing and Consumption during COVID-19 Pandemic and Possible Mitigation Strategies: A Bangladesh Perspective

**DOI:** 10.1101/2020.10.09.20210377

**Authors:** Monira Parveen, Mahmuda Yeasmin, Md. Maruf Ahmed Molla

**Affiliations:** Lecturer, Department of Pharmacology, Dhaka Dental College, Dhaka, Bangladesh; Medical Officer, Department of Virology, National Institute of Laboratory Medicine and Referral Center, Dhaka, Bangladesh

**Keywords:** COVID-19, anti-microbial resistance, AMR, Bangladesh

## Abstract

There are evidences that show increased antimicrobial consumption among COVID-19 patients. This has increased the burden on worsening situation of antimicrobial resistance (AMR) throughout the world. Bangladesh, one of the countries with highest numbers of COVID-19 cases, without effective regulation of antimicrobial prescription may suffer in future with study results showing a significant proportion of participants taking antimicrobial without proper indication and prescription from physicians. Suggested mitigation strategies include – strict regulation of over the counter (OTC) antimicrobial prescription, testing biochemical marker such as procalcitonin prior to initiation of antimicrobial therapy, introduction of color coded and tightly sealed bottled antimicrobial drugs, massive campaigning on social media, effective utilization of telemedicine and finally, raising awareness among physicians and patients regarding judicial use of antimicrobial.

## Introduction

COVID-19, an ongoing pandemic, has already created immense burden on health and social care system worldwide^1, 2^. One of the major issues encountered by physicians and researchers are lack of effective and safe treatment options against COVID-19. Hence, a plethora of anti-microbial agents and biological medicines have already been used either on a trial basis or as a last gasp measure to save patient lives. But, increased prescribing of antimicrobial, without first validating their effectiveness in treating COVID-19 cases, might flare up antimicrobial resistance (AMR) among patients^3^. On the other hand, increased societal awareness towards threats from infectious diseases, rational prescribing of antimicrobial drugs and good sanitary practices during this pandemic may subsequently have beneficial effect on AMR^4^. This short report will discuss issues of AMR during this era of COVID-19 including, but not limited to, how the ongoing pandemic might complicate the ongoing fight against AMR, evidences of indiscriminate antimicrobial usage among Bangladeshi COVID-19 patients, and how developing countries like Bangladesh should react to fight off this looming threat of AMR.

### COVID-19 and AMR: Fighting on two fronts

Around 700000 people die each year due to antibiotic resistance. If urgent measures are not taken, by the year 2050 around 10 million people would lose their lives to resistant microbes culminating in a financial loss similar to that of 2008-09 global financial crisis^5^. This projection is applicable for all countries irrespective of financial prowess, but the burden would be felt heavier by low-middle and low income countries including densely populated countries in South Asia and parts of Africa^5^.

The main challenge to manage patients of COVID-19 is its wide and unpredictable range of clinical manifestations. Around 84% people suffering from COVID-19 have mild or uncomplicated symptoms^6^. On the other hand, 14% may develop severe illness and need to be hospitalized, and among them 5% require ICU management^6^. However, only 1 to 10% of patients have been reported to contact secondary bacterial infections^7^. Despite the low rate of secondary bacterial infections, 45% of COVID-19 patients are receiving antibiotic therapy irrespective of their disease severity^8^.

The widespread usage of multi-drugs among hospitalized and critically ill patients may also increase AMR burden^3^. Literature evidences regarding bacterial and fungal co-infection among hospitalized patients are currently lacking, but nevertheless, a significant proportion of them receive multidrug and broad spectrum antibiotic therapy, rendering them vulnerable against selection of multidrug resistant (MDR) organisms^9^.

Azithromycin (in the macrolide and ketolide class), as well as vancomycin, carbapenems, tigecycline, ceftriaxone and linezolid, which are all classified as critically important antimicrobials (CIA) by WHO are being widely prescribed during this pandemic^10^. WHO also reports that, though combination of azithromycin and hydroxychloroquine is not recommended outside clinical trials, combination drugs are being used randomly at different hospitals around the world^11^. Moreover, sudden collapse of patients triggering stress and anxiety, with unavailable expert opinion on antibiotic usage may lead towards inappropriate or overuse of antibiotics^12^. Furthermore, a very common misconception among the public is that antibiotics can be used for viral infections (i.e., the common cold)^13^, and in developing countries like Bangladesh and India, where buying antibiotics from pharmacy is a common practice^14^, COVID-19 may further complicate the ongoing fight against AMR.

To prevent high infection rate, antimicrobial soaps and disinfectant cleaners are used widely as the most effective measure against COVID-19 infection, which can also have negative impact on AMR as they contain biocides that are antimicrobials^15^ This exposure of biocidal agents may increase the risk of cross resistance to antibiotics^16^, particularly those that treat Gram-negative bacteria^17^.

Here, the use of antimicrobials among COVID-19 patients gives rise to a dilemma, as without any recognized therapeutic agent to treat COVID-19 or current unavailability of a safe vaccine, physicians around the world are being forced to choose a wide range of antimicrobials in a desperate attempt to halt the progression of disease or exacerbation of a secondary condition in an individual. So, there are ultimately two different challenges to combat here – one is to treat COVID-19 patients, and another challenge is not to increase the burden of AMR by prescribing broad spectrum and unnecessary antimicrobial drugs.

### Irrational prescribing and overuse of anti-microbial during COVID-19: Evidences from Bangladesh

There is a well-established national guideline for clinical management of COVID-19 patients in Bangladesh. Without any sign of significant secondary bacterial infection, irrational antibiotic prescribing is prohibited within the national guideline^18^. Despite repeated attempts to avert potential complicated situation like AMR, preliminary reports from national newspaper and hospital records suggest COVID-19 patients with mild symptoms are usually found to be taking self-prescribed antimicrobials such as macrolides (azithromycin), doxycycline, anti-malarial drug (hydroxychloroquine), anti-parasitic (Ivermectin), and even trial drugs such as Favipiravir and Remdesivir without consulting physicians and completely disregarding their potential harmful side-effects^19^.

In order to further evaluate the extent of irrational antimicrobial usage, 100 covid-19 PCR positive patients, selected randomly from a tertiary COVID-19 PCR testing center in Dhaka, were contacted over phone and details regarding their sign-symptoms, drug history and consultation with physician/hospital admission history were sought. Among those participants, 96 of them gave accounts of symptoms classified as “mild disease” under the national guideline for COVID-19 clinical management, with most study participants (n=81) citing mild to moderate fever as one of their presenting complaints before opting for SARS-COV-2 PCR. Other notable symptoms included cough/sneezing/runny nose/ sore throat (n=32), headache (n=19), muscle ache (n=15), anosmia (n=8), chest pain/respiratory distress (n=4), diarrhea (n=3), and finally in a number of cases there were no symptoms (n=18).

Participants were asked questions regarding their health service seeking behavior (consultation with a physician) and their drug history. Surprisingly, only 55 participants consulted with a physician after being diagnosed as COVID-19 positive, while remaining 45 participants did not consult any physician/ healthcare worker after being diagnosed.

Detailed drug history were sought from participants with mild COVID-19 symptoms. A significant number of them (n=61) had one or more anti-microbial in their treatment regimen. Azithromycin was the most frequently prescribed drug (n=46) followed by ivermectin (n=22), doxycycline (n=21), and fluoroquinolones such as levofloxacin (n=6), moxifloxacin (n=4), and anti-viral drug such as favipiravir (n=4). Interestingly, only 25 of them had single anti-microbial agent in their treatment regimen with remaining 36 participants taking two or more anti-microbial simultaneously after being diagnosed as COVID-19 positive.

**Table-1:** Data regarding disease severity, health service seeking behavior, and antimicrobial prescription among study participants

|  | Frequency |
| --- | --- |
| **Disease Severity (n=100)** |  |
| Mild | 96 |
| Moderate/Severe/ Critically ill | 4 |
| **Presenting Symptom (n=100)** |  |
| Fever | 81 |
| Cough/ Sneezing/ Runny nose | 32 |
| Headache | 19 |
| Muscle ache | 15 |
| Chest pain/respiratory distress | 9 |
| Anosmia | 8 |
| Diarrhea | 3 |
| No symptom | 18 |
| **Health service seeking behavior (n=100)** |  |
| Consult with physician/Health care worker | 55 |
| No consultation | 45 |
| **Antimicrobial prescription (n=61)** |  |
| Treated with single agent | 25 |
| Combination of more than one antimicrobial | 36 |
| **Prescribed Antimicrobial drugs (n=61)** |  |
| Azithromycin | 46 |
| Ivermectin | 22 |
| Doxycycline | 21 |
| Levofloxacin | 6 |
| Moxifloxacin | 4 |
| Favipiravir | 4 |

### Mitigation Strategies to counteract potential increase in AMR

The intervention and mitigation strategies should include strict implementation of clinical treatment guidelines of COVID-19 to limit unnecessary antimicrobial usage, appropriate measures to intensify routine surveillance of AMR and restriction of over-the counter (OTC) sales of antimicrobial by law enforcement^20^. Ensuring appropriate antimicrobial in hour of need while avoiding unnecessary or prolonged antimicrobial course should be the principle of antimicrobial prescribing^21^.

Obtaining adequate microbiological evidence of pathogen before prescribing antimicrobial is of paramount importance to overcome this challenge. Unfortunately, this can be time consuming since there is a lack of advanced laboratory facilities within the country, and hence often puts the physician in a dilemma, especially when patient is critically ill and requires urgent intervention. As a result, without proper dosing and indications nearly half of all antimicrobial are being prescribed, which may contribute to the development of resistant pathogens^22^.

In order to overcome this issue, procalcitonin, an important serum biochemical marker can serve as a helpful adjunct to clinical judgment to distinguish between bacterial and viral infection especially in Lower Respiratory Tract Infection (LRTI) and for guiding antibiotic therapy as it remains undetectable in healthy individual^23^. Bacterial endotoxins as well as inflammatory cytokines such as TNF, IL-1B, and IL-6 act as a trigger for increased production of procalcitonin^24, 25^.

Little is known about correlation between SARS-CoV-2 and serum level of procalcitonin. However, rise of serum procalcitonin level in COVID-19 patients was observed in patients who required hospitalization or ICU admission explaining plausible bacterial co-infection or release of cytokine due to severe inflammatory response^26, 27^. In this regard procalcitonin might act as a guiding tool to initiate antibiotic in COVID-19 infected patients or patients presented with symptoms of LRTI during this pandemic.

There should be a clamping down on irrational prescription of anti-microbial without proper advices from clinicians. One possible solution in Bangladeshi context could be introduction of color coded strips of anti-microbial so that people purchasing them from pharmacy might know the potential harms associated with consumption these drugs without proper indication^28^.

Another reason behind anti-microbial resistance is not taking drugs for suggested duration treatment. People usually purchase anti-microbial from pharmacy and as soon as symptoms are alleviated, they stop taking anti-microbial without completing the course. This might be solved with introduction of tightly sealed bottle containing anti-microbial drugs sufficient for a 3, 7, 14 or 21 day course.

Social media platforms may play influential role in educating and raising awareness among antimicrobial prescribers, patients, and the general public on the significance of antibiotics in viral infections. Visual aids such as posters, brochures, and advertisements could be used in the local media as worthwhile intervention strategies. Awareness campaign displaying local surveillance data and sharing real life experience of individuals who have suffered from COVID-19 or any superbug in the past may help create a positive impact within the communities^29^.

Motivating physicians to start communicating with patients about the judicial use of antimicrobial and negative effect of AMR on health and economy might significantly improve the chances of success of such campaign^30^. Besides these, telemedicine could play potential role in this regard. This innovative technology allows physicians to provide critical support to COVID-19 patients as well as routine services to general people including valuable lessons on AMR^31^.

## Conclusion

The report illustrated the central importance of rational prescribing of anti-microbial during COVID-19 pandemic. It is evident that, Bangladesh, with a fragile health infrastructure, is currently struggling to curb down on overuse of antimicrobial among COVID-19 patients with a significant percentage of them, mostly with mild disease, taking one or more anti-microbial without consultation from physician or health care worker. More and extensive researches are recommended in near future to understand the true nature of antimicrobial overuse among Bangladeshi COVID-19 patients.

## Data Availability

All data regarding participants are available and could be provided on request

## Ethical clearance

Ethical clearance for this study was taken from ethical review committee of Dhaka Dental College (DDC/2020/42) on July 5, 2020.

